## Supplementary figures and images for "Exploring the molecular basis of the genetic correlation between body mass index and brain morphological traits"

### Supplementary Figure 1

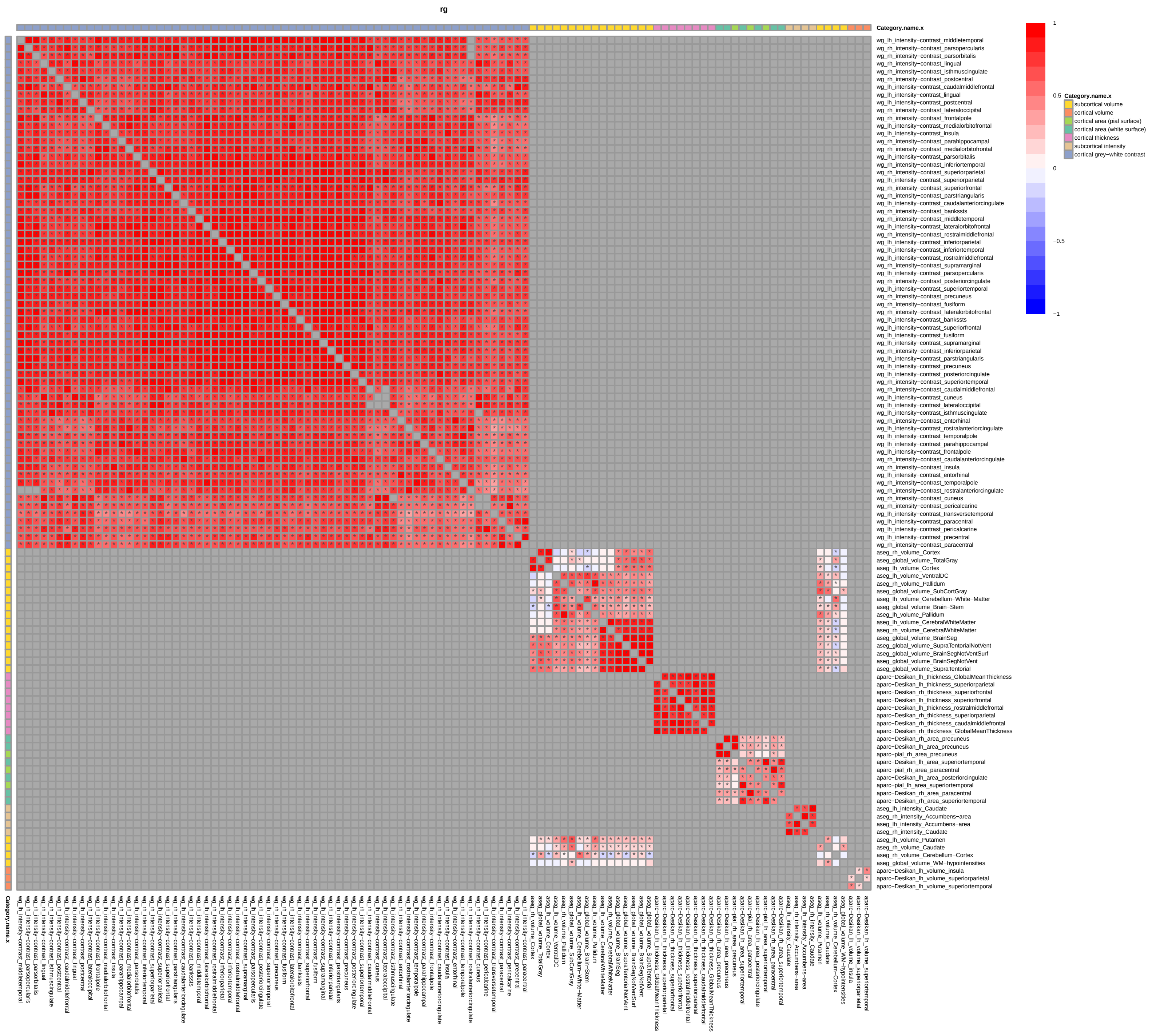

### Supplementary Figure 2

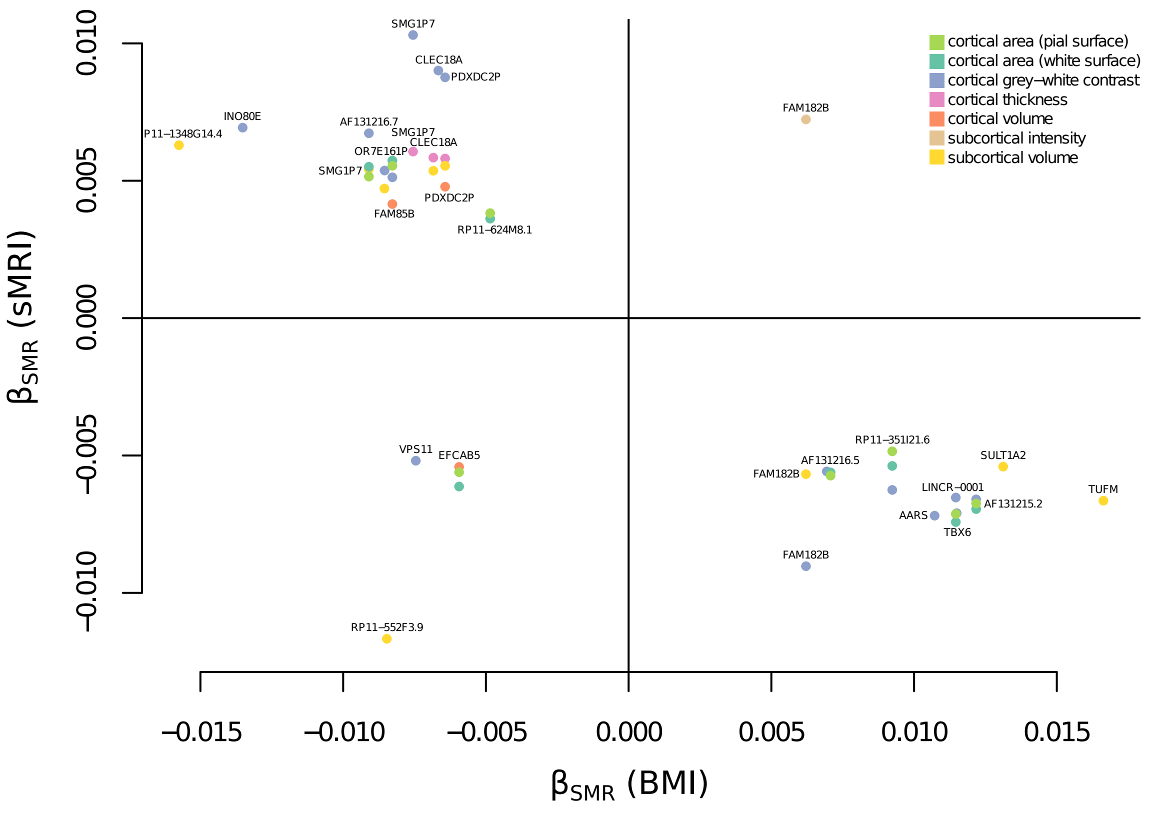

### Supplementary Figure 3

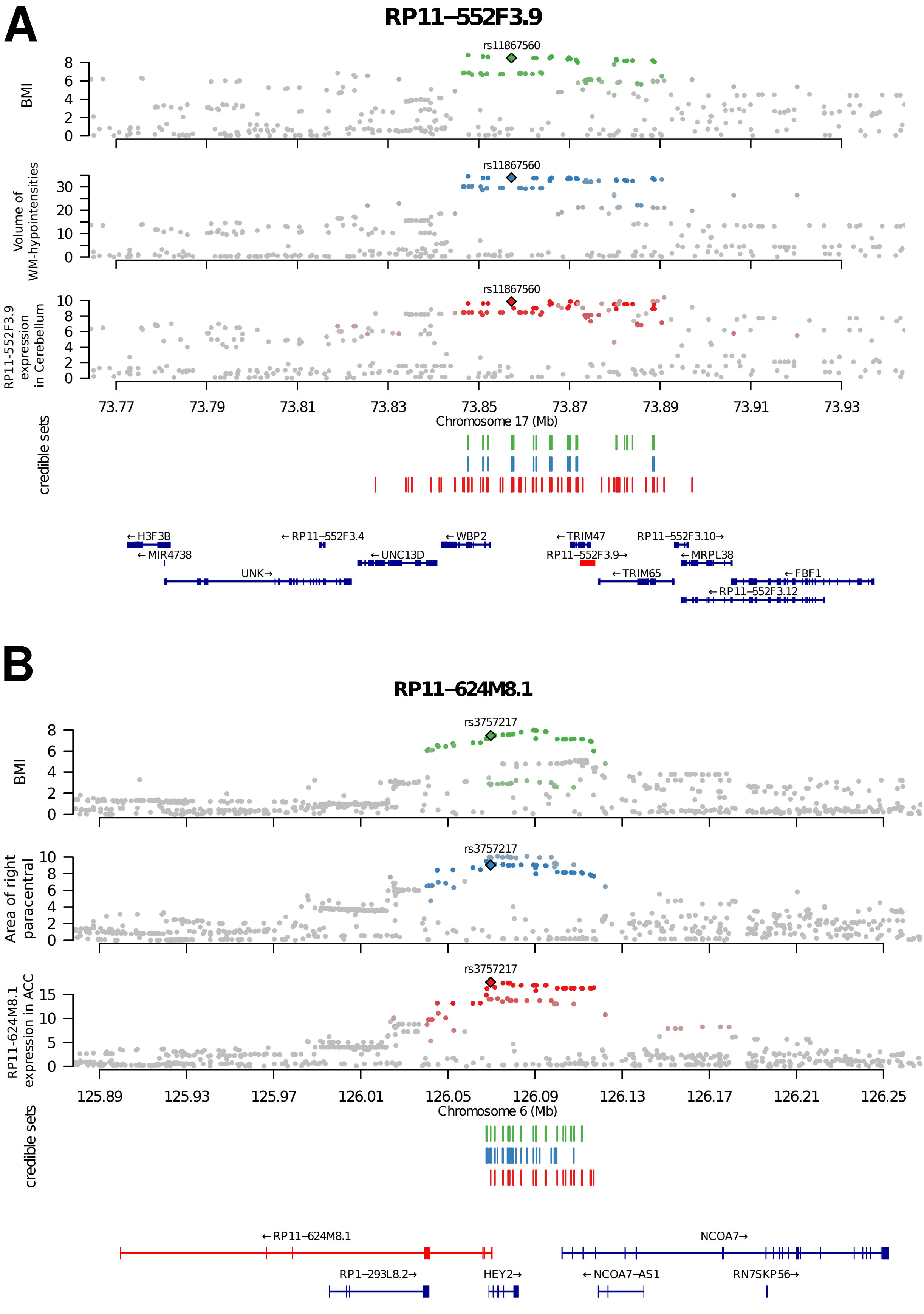
